# COVID-19 Hospitalizations in Five California Hospitals

**DOI:** 10.1101/2021.01.29.21250788

**Authors:** Miriam Nuño, Yury García, Ganesh Rajasekar, Diego Pinheiro, Alec J. Schmidt

**Affiliations:** Department of Public Health Sciences, University of California, Davis, United States; Department of Surgery, University of California, Davis, United States; Department of Internal Medicine, School of Medicine, University of California, Davis; Centro de Investigación en Matemática Pura y Aplicada (CIMPA), University of Costa Rica, San José, CR

**Author notes:** Corresponding author: Miriam Nuño, PhD; Department of Public Health Sciences, One Shields Avenue, Medical Sciences 1C, Davis CA 95616, United States. Reprints will not be available from the authors.

**Keywords:** severe acute respiratory syndrome coronavirus 2, coronavirus disease 2019, hospitalization, comorbidities, intensive care unit, in-hospital mortality, composite outcome

## Abstract

**Importance:** Characterization of a diverse cohort hospitalized with COVID-19 in a health care system in California is needed to further understand the impact of SARS-CoV-2 and improve patient outcomes.

**Objectives:** To investigate the characteristics of patients hospitalized with COVID-19 and assess factors associated with poor outcomes.

**Design:** Patient-level retrospective cohort study

**Setting:** University of California five academic hospitals.

**Participants:** Patients ≥18 years old with a confirmed test result for SAR-CoV-2 virus hospitalized at five UC hospitals.

**Exposure:** Confirmed severe acute respiratory syndrome coronavirus 2 (SARS-CoV-2) infection by positive results on polymerase chain reaction testing of a nasopharyngeal sample among patients requiring hospital admission.

**Main Outcomes and Measures:** Admission to the intensive care unit, death during hospitalization, and the composite of both outcomes.

**Results:** Outcomes were assessed for 4,730 patients who were discharged or died during a hospitalization. A total of 846 patients were treated at UC Davis, 1,564 UC Irvine, 1,283 UC Los Angeles, 471 UC San Diego, and 566 UC San Francisco. More than 20% of patients were ≥75 years of age (75-84: 12.3%, ≥85: 10.5%), male (56.5%), Hispanic/Latino (45.7%), and Asian (10.3%). The most common comorbidities were hypertension (35.2%), cardiac disease (33.3%), and diabetes (24.0%). The ICU admission rate was 25.2% (1194/4730), with 7.0% (329/4730) in-hospital mortality. Among patients admitted to the ICU, 18.8% (225/1194) died; 2.9% (104/3536) died without ICU admission. The rate of the composite outcome (ICU admission and/or death) was 27.4% (1,298/4,730). While controlling for comorbidities, patients of age 75-84 (OR 1.47, 95% CI: 1.11-1.93) and 85-59 (OR 1.39, 95% CI: 1.04-1.87) were more likely to experience a composite outcome than 18-34 year-olds. Males (OR 1.39, 95% CI: 1.21-1.59), and patients identifying as Hispanic/Latino (OR 1.35, 95% CI: 1.14-1.61), and Asian (OR 1.43, 95% CI: 1.23-1.82), were also more likely to experience a composite outcome than White. Patients with 5 or more comorbidities were exceedingly likely to experience a composite outcome (OR 2.74, 95% CI: 2.32-3.25).

**Conclusions:** Males, older patients, those with pre-existing comorbidities, and those identifying as Hispanic/Latino or Asian experienced an increased risk of ICU admission and/or death.

**KEY POINTS:** *Question:* What are the characteristics and outcomes of patients with SARS-CoV-2 infection hospitalized at five UC Health medical centers in California?

*Findings:* In this retrospective case series of 4,730 patients requiring hospitalization for COVID-19 in UC Health’s five medical centers, male (OR 1.41, 95% CI: 1.23-1.61), Hispanic/Latino (OR 1.35, 95% CI: 1.14-1.61), and Asian (OR 1.43, 95% CI: 1.12-1.82) were more likely to be admitted to the ICU and/or die after adjustment for age and comorbidity. ICU admission and/or death was more likely among older individuals and greater numbers of pre-existing conditions.

*Meaning:* This study describes the experience of a large, diverse cohort of patients with COVID-19 hospitalized in five hospitals in California between December 14, 2019 and January 6, 2021.

## INTRODUCTION

Severe acute respiratory syndrome coronavirus 2 (SARS-CoV-2), the virus causing coronavirus disease 2019 (COVID-19) has been a devastating pandemic, with over 91 million confirmed cases and nearly 2 million deaths worldwide.^1^ In the United States, the state of California (hereafter referred to as CA) leads the charts for the worse surge across the nation, with a total of 70,561 confirmed cases per million residents, 1,176 confirmed deaths per million residents, and potentially 1,173 ICU beds available for a population of nearly 40 million (January 12, 2021). Advanced age, chronic underlying conditions (including hypertension, diabetes, chronic kidney disease, among others), and being male are associated with an increased risk of hospitalization and death from COVID-19.^2-14^ There is additional, mounting evidence that some racial and ethnic minority groups are at disproportionate risk.^3,5,6,11^ Nation-wide, the Black/African American, Hispanic/Latino, Asian, and American Indian/Alaskan Native (AIAN) populations have seen elevated crude rates of infection, hospitalization, and/or mortality compared to the White population.^15^ In trends similar to other states, estimates from CA indicate that the Hispanic/Latino population make up 55% of confirmed cases and 47% of deaths and only 39% of the total population the Black/African American population represent 4% of confirmed cases and 7% of deaths and only 6% of the total population; and the Asian population represent 7% of all cases but 12% of all deaths. Multiple informal analyses report that Asians have low rates of confirmed cases but concerningly high case fatality rates,^16^ but few studies have been large enough to capture associated risk factors in this population.

More diverse, localized information is needed about who is being hospitalized with COVID-19 and their outcomes in the United States, and California in particular, especially since the second surge in the last half of 2020. This study describes the demographic characteristics, baseline comorbidities, and outcomes of patients hospitalized with COVID-19 in five acute care medical centers in the University of California Health System.

## METHODS

### Data Collection and Cohort Identification

In this retrospective cohort study, we identified patients who were hospitalized with laboratory-confirmed SARS-CoV-2 infections at any of the five UC Health hospitals between December 13, 2019 and January 6, 2021 by extracting electronic medical records (EMRs) from The COVID Research Data Set (CORDS). The index date corresponds to the first confirmed COVID-19 diagnosis. A hospitalization was included if test confirmation occurred within 21 days or during an inpatient admission. SARS-CoV-2 positive status was determined by PCR or SNOMED code 840539006 (Disease caused by Severe acute respiratory syndrome coronavirus 2) during their earliest hospitalization (**Supplemental eTable 1**).

Patient characteristics, including race/ethnicity, sex, comorbidities, and clinical outcomes were collected. Data was queried on January 8, 2021 and tabulated by January 10, 2021. We captured comorbidities using the *International Statistical Classification of Diseases and Related Health Problems, Tenth Revision* (*ICD*-*10*) classification (**Supplemental eTable 2**). Self-identified race/ethnicity was categorized into White, Hispanic/Latino, Black/African American, and Asian; Other includes AIAN (n=9, 0.19%), Native Hawaiian or Other Pacific Islander (n=53, 1.12%), Other (n=169, 3.57%), Unknown (n=246, 5.20%), and those that identified with multiple race/ethnicity (n=46, 0.97%).

Comorbidities included cancer, cardiac disease, cerebrovascular disease, coagulopathy, deficiency anemia, depression, diabetes, drug use disorders, HIV/Aids, hypertension, hypothyroidism, liver disease, neurological conditions, obesity, paralysis, pregnancy, psychoses, pulmonary disease, renal failure, collagen vascular disease, smoking status, and solid organ transplantation. Comorbidities documented 2 months prior to COVID-19 confirmation, during hospitalization, or 2 months post discharge were considered. The 2 month post-discharge observation period to capture comorbidities was deemed reasonable given lags in data entry due to billing. A comorbidity score represented the sum of any of these conditions. The lowest score of a 0 (n=1806, 38.2%) was assigned to patients without documented comorbidities. The co-occurrence of comorbidities was illustrated through correlation networks. The University of California Health System limited data set is de-identified and did not require IRB approval. The UC Davis implementation of the University of California COVID Research Data Set (CORDS) was determined exempt from human subject protection under IRB protocol 1604619-1.

### Statistical Analysis

Univariate and multivariable logistic regression was used to estimate the risk of admission to the ICU, in-hospital mortality, and the composite outcome of ICU admission and/or death during the hospitalization. The multivariable model adjusted for age in years at admission, sex, and race/ethnicity. Odds Ratios (OR) and 95% confidence intervals (95% CI) were reported.

We measured the strength of comorbidity associations with the Pearson’s correlation coefficient (*Φ*) for binary variables. The correlation coefficient between a pair of comorbidities, *Φ*_*ij*_, was calculated according to Equation [1] below.

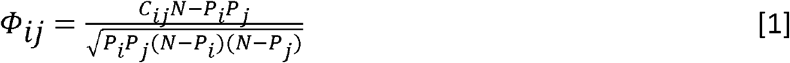

Where *C*_*ij*_ corresponds to the number of patients affected by both comorbidities, *N* is the total number of patients in the study, and *P*_*i*_ is the prevalence of the *i*^*th*^ comorbidity. The distribution of *Φ* values represent all disease pairs where *C*_*ij*_ > 0. The correlation for the dichotomous variable is denoted by *Φ* and a t-test is used to determine the significance of *Φ* ≠ 0.^17^ We visualized comorbidities with correlation (*Φ* > 0) larger than expected by chance for the entire cohort, as well as stratified by composite outcome.

We described patterns in cumulative hospitalizations during the study period over time by using change-point analysis to discern specific time points where abrupt changes in the mean occurred. This algorithm detects multiple points of change in time-ordered data through binary segmentation.^18-20^ First, we applied a change-point statistical test to the entire time series of hospitalization cases. Then, we split the series into two new ones when a point change was found. We repeated this procedure several times until there were no further significant change points. Statistical analyses was conducted using SAS software, version 9.4 (SAS Institute, Cary, NC, USA), *changepoint* package (binary segmentation algorithm) in R version 4.0.1 (R Core Team 2020), and Python. Data were analyzed from January 8 to January 20, 2021.

## RESULTS

A total of 4,730 hospitalized patients with positive SARS-CoV-2 assays were included (median age 61 years, IQR: 46-73) (**Table 1**). 56.4% were male; one patient was missing sex data. A total of 339 (7.2%) identified as Black/African American, 2,161 (45.7%) as Hispanic/Latino, 489 (10.3%) as Asian, 523 (11.1%) were documented as Other/Unknown, and 1,218 (25.8%) identified as White. A moderate fraction of patients did not have documented comorbidities (n=1,806, 38.2%). The most common comorbidities were hypertension (35.2%), cardiac disease (33.3%), diabetes (24.0%), pulmonary disease (17.8%), obesity (17.3%), and renal failure (15.3%). Coagulopathy (11.8%), smoking (10.2%), depression (9.4%), liver disease (8.8%), and cerebrovascular disease (7.8%) were also reported. Among the possible 22 comorbidities documented for each individual patient, we found that 457 (9.7%) patients had one, 488 (10.2%) had two, 468 (9.9%) had three, 430 (9.1%) had four, 355 (7.5%) had five, and 726 (15.4%) had six or more comorbidities.

**Table 1.**
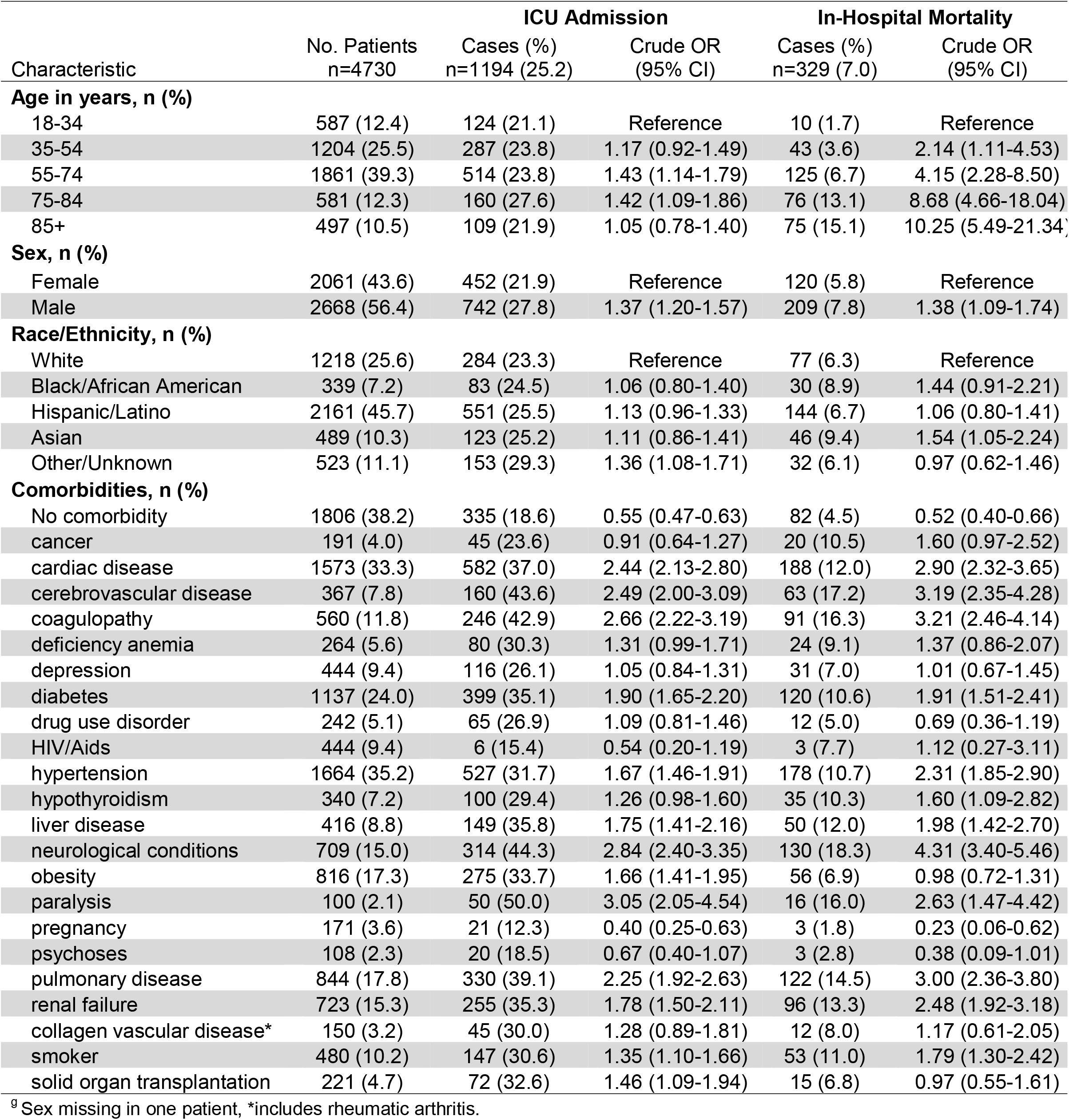
Characteristics and comorbidities of 4,730 patients with COVID-19 hospitalized in UC Health Hospitals.

### Quantifying the Strength of Comorbidity Relationships

We quantified the network of comorbidities and visualized correlations that were positive and significant by linking chronic conditions in a visual network (**Figure 1**). Figure 1A displays correlations across the total cohort, of which hypertension and heart disease showed the strongest correlation; Figure 1B displays the correlation network among patients that did not experience a composite outcome; and Figure 1C describes the correlation network of comorbidities for patients that experienced a composite outcome. Figures 1B and 1C together suggest that hypertension, diabetes, cardiac disease, renal failure, neurological disease, and depression, among other diseases, tend to be more comorbid in individuals that experienced a composite outcome than in those that did not. The comorbidity network structure in Figure 1A summarized and reflected well-known associations with COVID-19 hospitalizations, while suggesting that the interactions between comorbidities are expressed differently in those with more severe and deadly cases.

**Figure 1.**
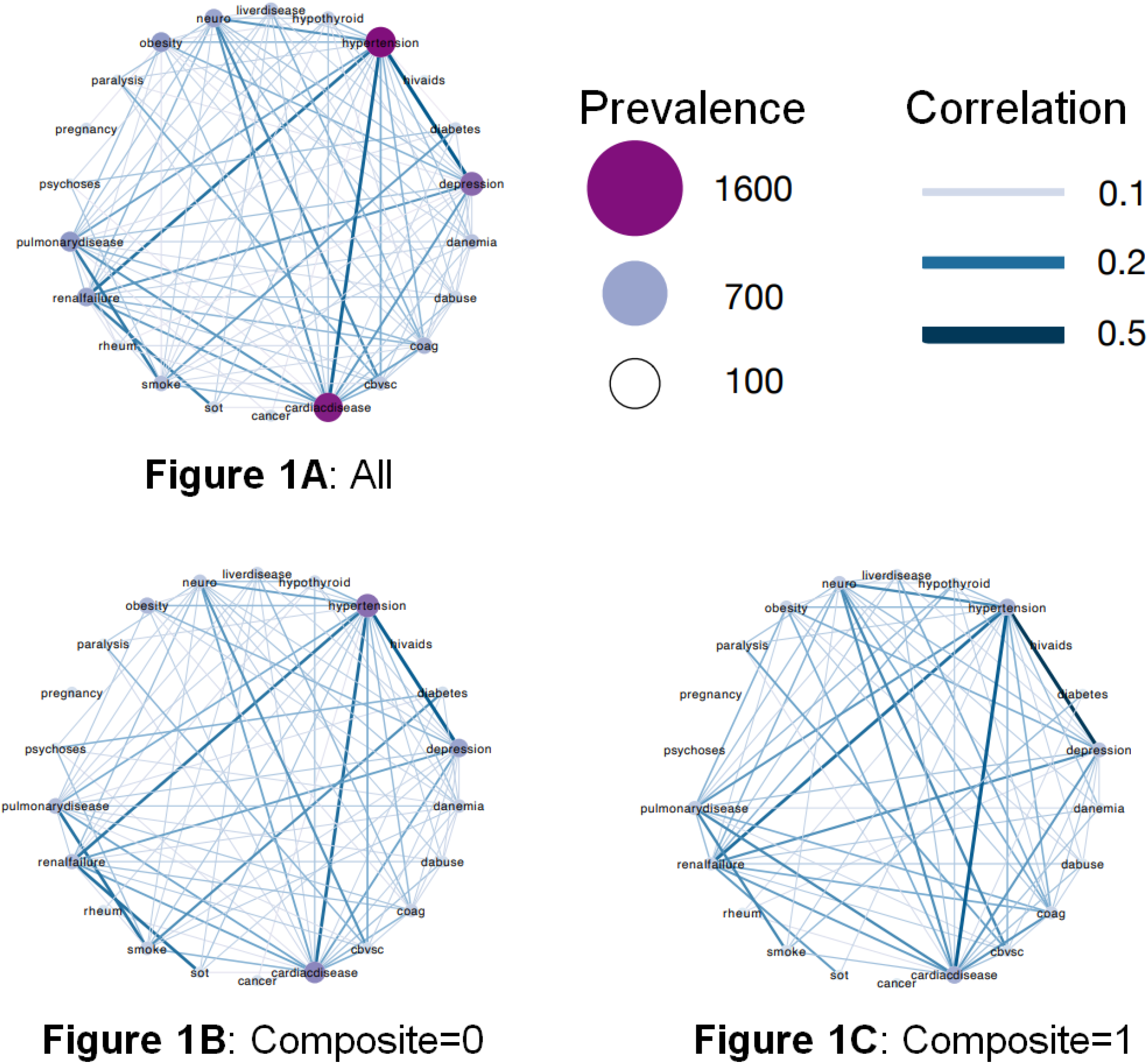
Comparison between the strength of comorbidities observed in patients overall, those who experienced a composite outcome and those that did not. Network includes comorbidities with positive and significant correlation.

### Quantifying Changes in Hospitalization, ICU Admission and In-hospital Mortality

**Figure 2** shows the daily cumulative COVID-19-related hospital admissions, ICU admissions, and deaths during hospitalization. Change-point analysis showed that April 18, May 18, June 25, August 14, September 11, October 15, and November 21 corresponded to significant abrupt changes in the number of hospitalizations. Similar analysis for the number of ICU admissions and deaths are included in Supplemental Table 3. Similar abrupt changes in ICU admissions occurred on the same dates as those for hospitalization; death trends changed on April 15, May 26, July 3, August 6, September 13, October 16, and November 22, 2020.

**Figure 2.**
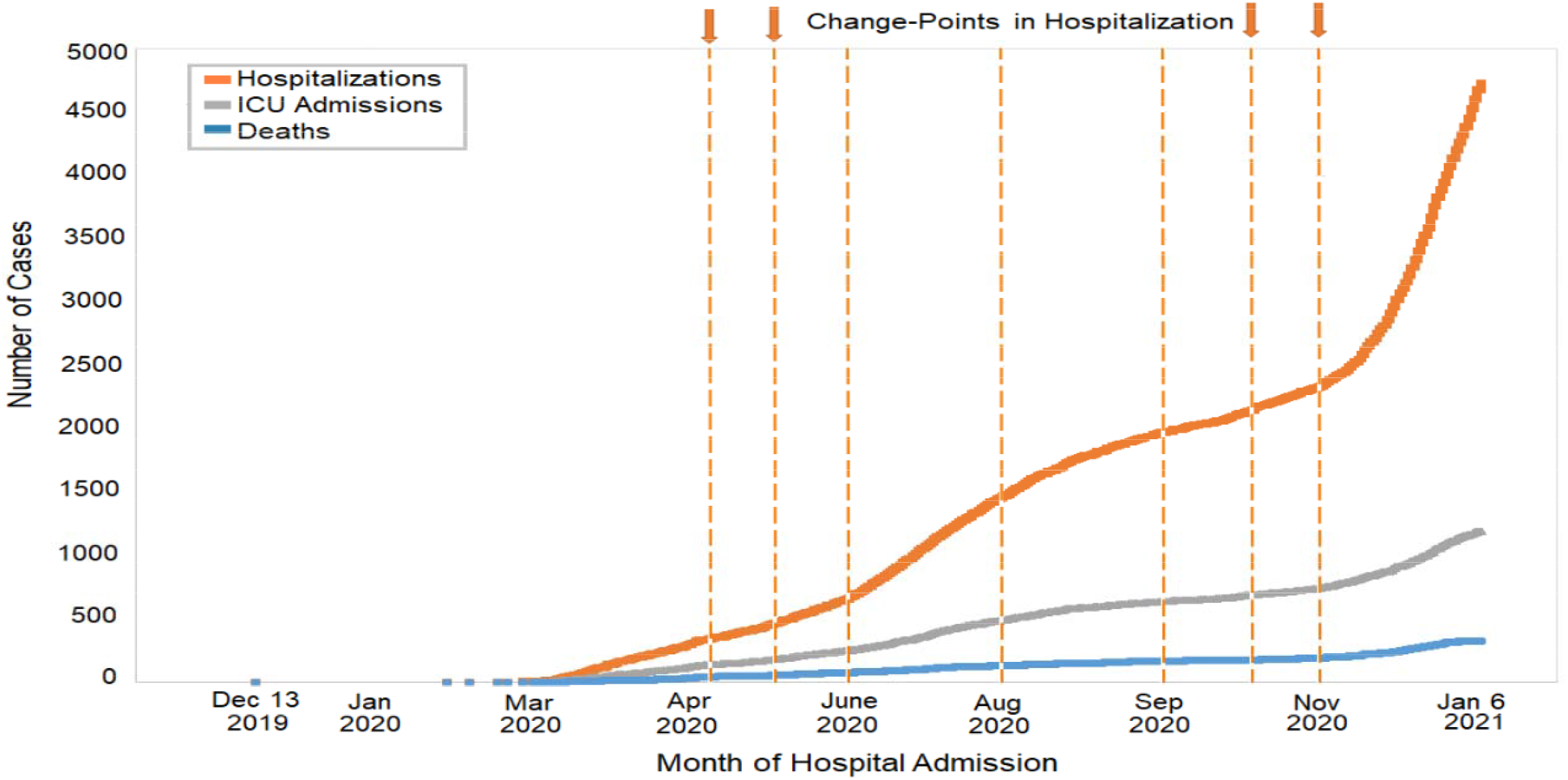
Cumulative Number of Patients with COVID-19 Hospitalized, admitted to the ICU, and Died in UC Health Hospitals, December 13, 2019 to January 6, 2021. Dashed vertical lines correspond to change points in hospitalization that occurred in April 18, May 18, June 25, August 14, September 11, October 15, and November 21, 2020.

### Univariable Analysis: ICU Admission

We identified 1,194 (25.2%) patients who were admitted to the intensive care unit with confirmed infection with SARS-CoV-2 (**Table 1**). In unadjusted univariate analyses, patients of age 55-74 and 75-84 years were more likely to be admitted to the ICU than 18-44 year-olds, and ICU admission was more likely in male patients than female. Hispanic/Latino, Black/African American, and Asian patients were no more or less likely to be admitted to the ICU once in the hospital than White patients, though Other races/ethnicities were (OR 1.36, 95% CI: 1.08-1.71). Among the common comorbidities, the most likely to be admitted to the ICU were those patients with neurological conditions (OR 2.84, 95% CI: 2.40-3.35), coagulopathy (OR 2.66, 95% CI: 2.22-3.19), and cerebrovascular disease (OR 2.49, 95% CI: 2.00-3.09). Other high-risk chronic conditions included cardiac disease, pulmonary disease, diabetes, hypertension, obesity, liver disease, and renal failure.

### Univariable Analysis: In-Hospital Deaths

A total of 329 (7.0%) died during hospitalization (**Table 1**). The risk of in-hospital mortality increased significantly with age, particularly among patients of age 85 years and older (OR 10.25, 95% CI: 5.49-21.34) compared to 18-34 year-olds. Male patients were more likely to die in-hospital than female (OR 1.38, 95% CI: 1.09-1.74). Hispanic/Latino, and Other patients were no more or less likely to die than White; however, Asians showed an increased likelihood of in-hospital mortality (OR 1.54, 95% CI: 1.05-2.24) and Black/African Americans were trending in the same direction (OR 1.44, 95% CI: 0.91-2.21), albeit with a smaller sample (n=46 vs. 31, respectively). The presence of 12 out of the 22 comorbidities investigated were significantly associated with increased odds of in-hospital mortality: neurological conditions (OR 4.31, 95% CI: 3.40-5.46), coagulopathy (OR 3.21, 95% CI: 2.46-4.14), and cerebrovascular disease (OR 3.19, 95% CI: 2.35-4.28), had the most pronounced association with in-hospital mortality. Diabetes, hypertension, liver disease, pulmonary disease, renal failure, and a current smoking history also increased the odds of in-hospital mortality.

### Multivariable Analysis: Composite Outcome

Overall, 27.4% (1,298/4,730) of patients requiring a hospitalization for COVID-19 were admitted in the ICU and/or died during their hospitalization, defined here as a “composite outcome.” The rate of patients with a composite outcome differed across the five UC Health Hospitals (UCD: 33.9%, 287/846; UCI: 18.2%, 285/1564; UCLA: 33.1%, 424/1283; UCSD: 22.9%, 108/471; UCSF: 34.2%, 194/566, p<0.001). Composite outcome rates also vary by month of hospital admission for 2020, with an outlier in March of 2020 at UCSF, due to a single hospitalization requiring ICU admission (**Figure 3**). In analysis adjusted for age and the number of relevant comorbidities, we found that older patients (85+ vs. 18-34, OR 1.39, 95% CI: 1.04-1.87), male (OR 1.41, 95% CI: 1.23-1.61), Hispanic/Latino (OR 1.35, 95% CI: 1.14-1.61), and Asian (OR 1.43, 95% CI: 1.13-1.82) were more likely to be admitted to the ICU and/or die during hospitalization (**Figure 4**). Patients with higher number of comorbidities were also more likely to experience a composite outcome (≥5 vs. none, OR 1.74, 95% CI: 2.32-3.25).

**Figure 3.**
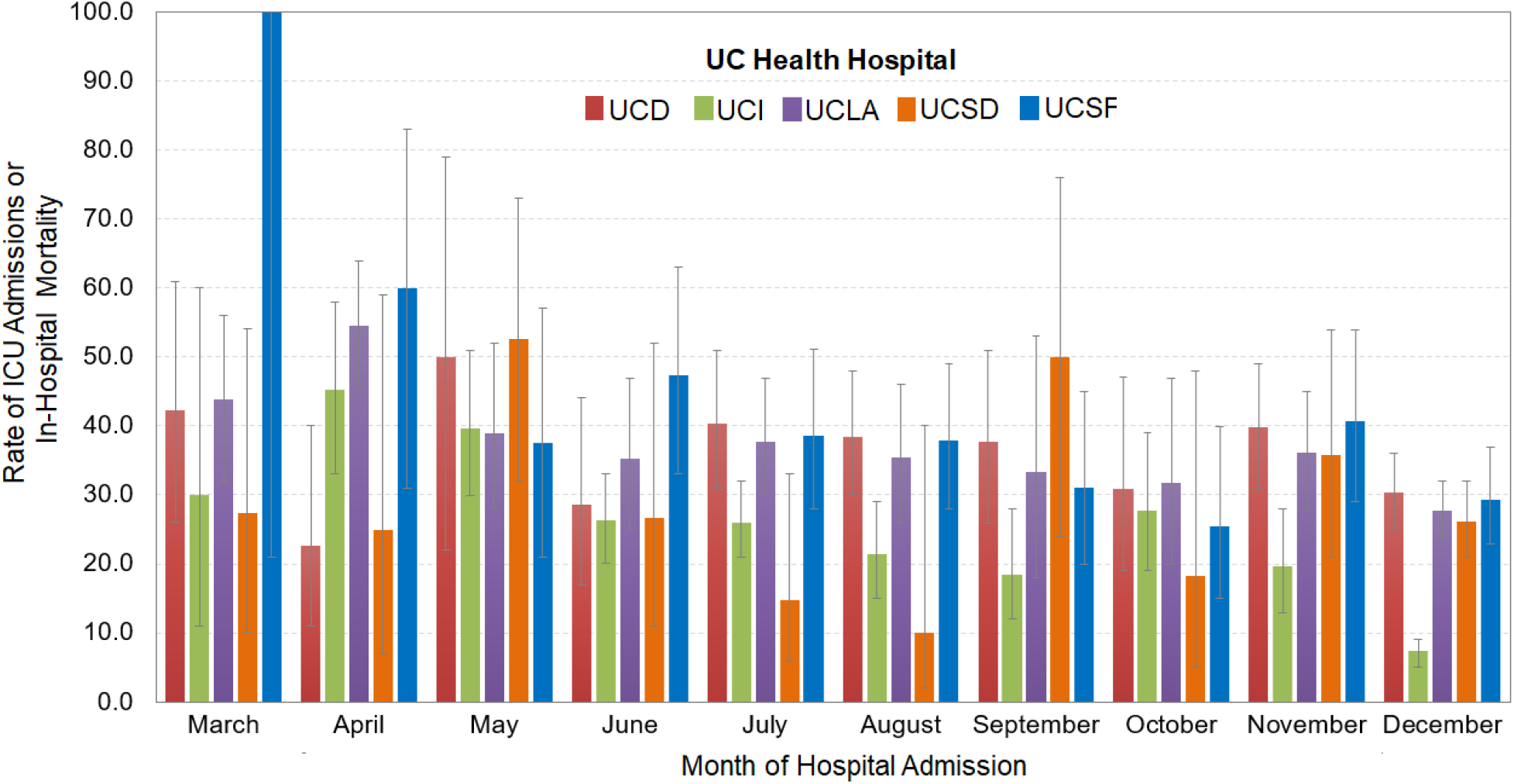
Rate (95% Confidence Intervals) of Patients Who Experienced a Composite Outcome of ICU Admission and/or Death, During Hospitalization for COVID-19 in UC Health Hospitals, 2020.

**Figure 4.**
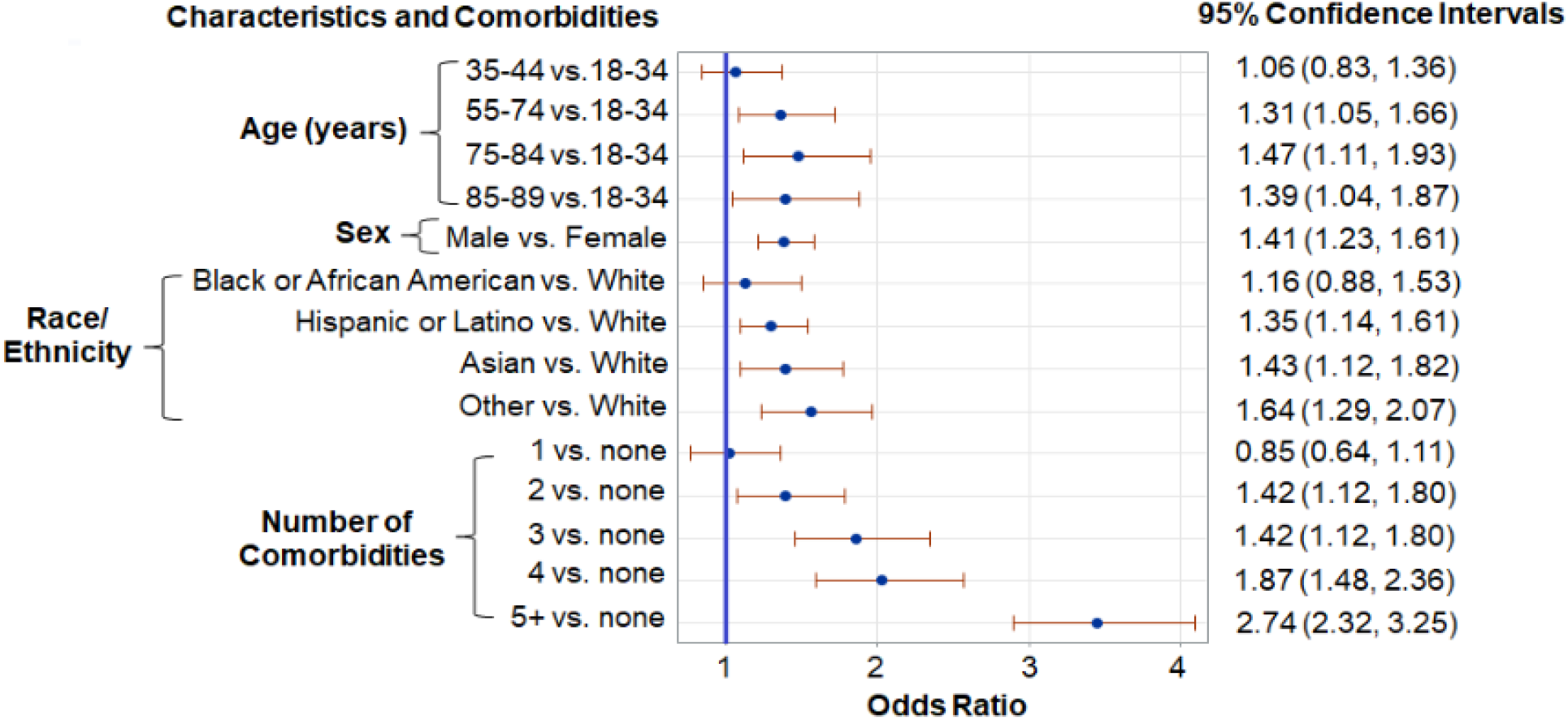
Multivariable Logistic Regression Results for Assessing Factors Associated with the Composite Outcome of In-Hospital Mortality and/or Admission to the ICU Among Patients Hospitalized for COVID-19 in UC Health Hospitals, December 2019 to January 2021.

## DISCUSSION

In this study of 4,730 patients with COVID-19 requiring hospitalization at five hospitals in California, 25.2% were admitted to the ICU, and 7.0% died. Patients were older (20% greater than 75 years), largely Hispanic/Latino (45.7%), with moderate representation of White (25.8%), Asian (10.3%), and Black/African American (7.2%). The 1-year cumulative ICU admission rate for COVID-19 is equitable to previous reports, but the in-hospital mortality rate is smaller than observed in case studies from earlier in course of the pandemic: ICU admission rates ranged from 14.2% to 35.7% in March-May 2020, where mortality rates ranged from 21.0% to 29.1% over the same period.^2,9,11,21^ Our study captured the experience of patients hospitalized for COVID-19 in five hospitals in California, across which the rates of composite outcome ranged from 18.2% to 34.2%. The variability of this outcome by hospital is likely influenced by differences in patient severity and treatment practices for COVID-19 across hospital settings, which have evolved throughout 2020, and regional fluctuations in the scope of the pandemic.

Differences observed in mortality and severity of COVID-19 among patients that require hospitalization are probably multifactorial. Our sample was drawn from a longer time period, which would include effects from improved treatment options and resource availability that were frequently lacking during earlier stages. Additionally, geographic differences in the distribution of races/ethnicities and the prevalence of chronic conditions that appear to increase the risk of severe illness may contribute to differences in hospital outcomes across the country. Criteria for hospital admission may have changed throughout this pandemic and likely will be driven, at least partially, by the availability of resources and ICU beds, which have fluctuated as well. Despite the documented differences in the outcomes of hospitalized COVID-19 patients, trends in these outcomes across the five hospitals during the study period are consistent, with reasonable fluctuations of the outcomes assessed.

The associations with older age, comorbidities, and race/ethnicity with poor outcomes linked to COVID-19 hospitalization are largely consistent with previous findings. This study also highlighted the increased risk of poor outcomes in the Asian population of California. In most states and counties, the relatively small proportion of Asian in the population makes awareness of cases and deaths in this community more challenging, as general samples large enough to make conclusions from are difficult to come by. California is one of few states with a sufficiently sizable Asian population for their COVID-19 burden to become unmistakable in the aggregate. There is growing recognition of the burden of COVID-19 among Asian, but data on outcomes among Asian ethnic subgroups remain extremely limited. A recent systematic review and meta-analysis of 50 studies from the US and United Kingdom found higher risk of COVID-19 infection among Asians and Blacks compared to Whites, and possibly an increased risk of ICU admission and death only among Asians compared to Whites.^22^ Further studies are needed to investigate the impact of COVID-19 infection and severity in Asian populations, particularly addressing potential differences in Asian ethnic subgroups at a broader geographic scale. These investigations will be critical to appropriately allocate resources to hardest hit communities, including testing and communication regarding seeking care as well as public health policies to mitigate risk factors and improve health equity.

The composite outcome in this study involved an ICU admission and/or in-hospital mortality, which occurred in 27.5% of patients. We reported rates only for patients who were discharged alive or died during the hospitalization by the end of the study period. 61.8% of patients had 1 or more relevant comorbidities and nearly 23% had 5 or more of these pre-existing conditions, which substantially increased the odds of a poor outcome. Using a network-based approach, we found that hypertension, cardiac disease, diabetes, and renal failure are positively and significantly associated to each other in the overall population, a relationship even more evident in patients with a composite outcome.

This study has several limitations. Our findings represent the experience of five hospitals within the UC Health system and therefore may have limited external generalizability to other health care settings, especially outside of California. Since our sample was drawn from electronic medical records and not full chart reviews, our data was also missing reported symptoms and the patients’ stated reasons for seeking COVID-19 tests, which could be one or a combination of the presence of COVID-19 symptoms, notification of exposure through contact tracing, or seeking a test in lieu of observing a 14-day quarantine period after travel. Such a large cohort followed over an extended time of period will necessarily include aggregation biases due to differential test availability, hospital admission practices, and documentation across the five medical centers, especially as public health advice evolved over the observation period. This could very well have obscured evolving demographic trends as the pandemic developed, or introduced a confounding bias in crude measures. Finally, cases who were tested at UC Health and sought care outside the system were not captured in this database, meaning any subsequent mortality or ICU admission of these cases outside the UC Health system would not be captured either. Notwithstanding these limitations, this study provides comparative epidemiologic characteristics of a diverse and underrepresented population of patients admitted to the hospital with COVID-19 over an entire year of the pandemic. Particularly relevant is the captured experience of Asians hospitalized due to COVID-19, and the differences in demographic characteristics and associations of comorbidities with each other and with poor outcomes.

## Conclusions

In this study, we found that older patients, with multiple comorbidities, identifying as Hispanic/Latino or Asian were more likely to be admitted to the intensive care unit and/or die during hospitalization at five UC Health medical centers. The overall mortality rate observed is significantly lower than what has been documented in hospitalized patients with COVID-19 from the early stages of the pandemic, despite the reported high levels of comorbidities. These findings provide additional evidence of the impact of COVID-19 in system of hospitals in California. While the comparatively low mortality rate is reassuring, the differential impact on racial/ethnic minorities requires the implementation of an equitable solution.

## Data Availability

CORDS is de-identified but is still protected health information under HIPAA, and is therefore not publicly available.

## ABBREVIATIONS

SARS-CoV-2 =: Severe Acute Respiratory Syndrome Coronavirus 2
COVID-19 =: coronavirus disease 2019
UC Health =: University of California Five Academic Medical Centers
UCD =: University of California Davis, Medical Center
UCI =: University of California Irvine, Medical Center
UCLA =: University of California Los Angeles, Medical Center
UCSD =: University of California San Diego, Medical Center
UCSF =: University of California San Francisco, Medical Center
ICU =: Intensive Care Unit
ICU Days =: Days spent in the Intensive Care Unit
LOS =: Days spent in the hospital before discharge or death
ICD-10-CM =: International Classification of Diseases, 10^th^ Revision, Clinical Modification
AIAN =: American Indian/Alaskan Native

## ACKNOWLEDGEMENTS

Dr. Nuño wish to thank Dr. James M. Bugni and Blanca Nuño for supporting this research effort and providing critical feedback to improve this study. We additionally wish to thank members of the IT Health Informatics division at UC Davis. Hemanth Tatiparthi and Ranjit Singh created the local CORDS database, Steve Covington provided advice about querying the database, and Brian Paciotti helped evaluate data irregularities among sites.

## DECLARATION OF CONFLICTING INTERESTS

The authors declare that there is no conflict of interest.

## FUNDING/FINANCIAL SUPPORT

None Reported

## DISCLAIMER

The Limited Dataset used for this study

## AUTHOR CONTRIBUTIONS

Dr. Nuño had full access to the limited dataset in the study and takes full responsibility for the integrity of the data and accuracy of the analysis.

Concept and Design: Nuño, Rajasekar.

Acquisition, analysis, or interpretation: All Authors.

Drafting of Manuscript: Nuño, Schmidt.

Statistical Analysis: Nuño, García, Pinheiro.

## Notes

### Competing Interest Statement

The authors have declared no competing interest.

### Funding Statement

No external funding was received.

### Author Declarations

The University of California Health System determined that the COVID Research Data Set (CORDS), a de-identified and limited data set, was exempt from human subject protection under IRB protocol 1604619-1.

