## Supplementary Materials for "COVID-19 Hospitalizations in Five California Hospitals"

### Supplemental Online Content

**eTable 1**. Severe Acute Respiratory Syndrome Coronavirus 2 Lab Tests.
**eTable 2**. International Classification of Disease Codes for Comorbidities.
**eAppendix** 1. Correlation Network of comorbidities.
**eAppendix 2**. Change-Point Detection Analysis of Hospitalizations, ICU Admissions, and In-Hospital Deaths.
**eFigure 1**. Fits of data and change points

| **eTable 1. Severe Acute Respiratory Syndrome Coronavirus 2 Lab Tests.** | |
|---|---|
| Long Common Name | LOINC Code |
| SARS-CoV-2 (COVID-19) N gene [Presence] in Respiratory specimen by NAA with probe detection | 94533-7 |
| SARS-CoV-2 (COVID-19) RNA [Presence] in Unspecified specimen by NAA with probe detection | 94309-2 |
| SARS-CoV-2 (COVID-19) RNA [Presence] in Respiratory specimen by NAA with probe detection | 94500-6 |
| SARS-CoV-2 (COVID-19) RNA panel - Unspecified specimen by NAA with probe detection | 94306-8 |
| SARS-CoV-2 (COVID-19) RNA panel - Respiratory specimen by NAA with probe detection | 94531-1 |
| SARS-CoV-2 (COVID-19) RdRp gene [Presence] in Respiratory specimen by NAA with probe detection | 94534-5 |
| SARS-like coronavirus N gene [Presence] in Unspecified specimen by NAA with probe detection | 94310-0 |

**eTable 2. International Classification of Disease Codes for Comorbidities**

| Comorbidity | ICD-10-CM Codes |
|---|---|
| Cancer | C00, C01, C02, C03, C04, C05, C06, C07, C08, C09, C10, C11, C12, C13, C14, C15, C16, C17, C18, C19, C20, C21, C22, C23, C24, C25, C26, C30, C31, C32, C33, C34, C37, C38, C39, C40, C41, C43, C45, C46, C47, C48, C49, C50, C51, C52, C53, C54, C55, C56, C57, C58, C60, C61, C62, C63, C64, C65, C66, C67, C68, C69, C70, C71, C72, C73, C74, C75, C76, C77, C78, C79, C80, C81, C82, C83, C84, C85, C86, C88, C90.0, C90.2, C91.0, C91.1, C91.5, C92.0, C92.1, C92.4, C92.5, C92.6, C92.8, C96, C97 |
| Cardiac Disease | A52.0, I05, I06, I07, I08, I09.1, I09.8, I09.9, I11.0, I13.0, I13.2, I25.10, I25.5, I34, I35, I36, I37, I38, I39, I42.0, I42.5, I42.6, I42.7, I42.8, I42.9, I43, I44.1, I44.2, I44.3, I45.6, I45.9, I47, I48, I49, I50, I70, I71, I73.1, I73.8, I73.9, I77.1, I79.0, I79.2, K55.1, K55.8, K55.9, P29.0, Q23.0, Q23.1, Q23.2, Q23.3, R00.0, R00.1, R00.8, T82.1, Z45.0, Z95.0, Z95.2, Z95.3, Z95.4, Z95.8, Z95.9 |
| Cerebrovascular Disease | G45, G46, H34.0, I60, I61, I62, I63, I64, I65, I66, I67, I68, I69 |
| Coagulopathy | D65, D66, D67, D68, D69.1, D69.3, D69.4, D69.5, D69.6 |
| Deficiency Anemia | D50.8, D50.9, D51, D52, D53 |
| Depression | F20.4, F31.3, F31.4, F31.5, F32, F33,F34.1, F41.2, F43.2 |
| Diabetes | E10.0, E10.1, E10.2, E10.3, E10.4, E10.5, E10.6, E10.7, E10.8, E10.9, E11.0, E11.1, E11.2, E11.3, E11.4, E11.5, E11.6, E11.7, E11.8, E11.9, E12.0, E12.1, E12.2, E12.3, E12.4, E12.5, E12.6, E12.7, E12.8, E12.9, E13.0, E13.1, E13.2, E13.3, E13.4, E13.5, E13.6, E13.7, E13.8, E13.9, E14.0, E14.1, E14.2, E14.3, E14.4, E14.5, E14.6, E14.7, E14.8, E14.9 |
| Drug Abuse | F11, F12, F13, F14, F15, F16, F18, F19, Z71.5, Z72.2 |
| HIV/AIDS | B20, B21, B22, B24 |
| Hypertension | I10, I11, I12, I13, I15 |
| Hypothyroidism | E00, E01, E02, E03, E89.0 |
| Liver Disease | B18, I85, I86.4, I98.2, K70, K71.1, K71.3, K71.4, K71.5, K71.7, K72, K73, K74, K76.0, K76.2, K76.3, K76.4, K76.5, K76.6, K76.7, K76.8, K76.9, Z94.4 |
| Neurological Conditions | F01.5, F03.9, G10, G11, G12, G13, G20, G21, G22, G25.4, G25.5, G30, G31.2, G31.8, G31.9, G32, G35, G36, G37, G40, G41, G93.1, G93.4, R47.0, R56 |

| | |
|---|---|
| Obesity | E66, Z68.3, Z68.4, E66.01, E66.2, Z68.4 |
| Paralysis | G04.1, G11.4, G80.1, G80.2, G81, G82, G83.0, G83.1, G83.2, G83.3, G83.4, G83.9 |
| Pregnancy | O00, O01, O02, O03, O04, O05, O06, O07, O08, O09,O10, O11, O12, O13, O14, O15, O16, O20, O21, O22, O23, O24, O25, O26, O27, O28, O29, O30, O31, O32, O33, O34, O35, O36, O37, O38, O39, O40, O41, O42, O43, O44, O45, O46, O47, O48, O60, O61, O62, O63, O64, O65, O66, O67, O68, O69, O70, O71, O72, O73, O74, O75, O76, O77, O80, O82, O85, O86, O87, O88, O89, O90, O91, O92, O94-O9A, Z34.00, Z34.01, Z34.02, Z34.03, Z34.90, Z34.91, Z34.92, Z34.93 |
| Psychoses | F20, F22, F23, F24, F25, F28, F29, F30.2, F31.2, F31.5 |
| Pulmonary Disease | E84.0, E84.11, E84.19, E84.9, I26, I27, I27.8, I27.9, I28.0, I28.8, I28.9, J40, J41, J42, J43, J44, J45, J45.4, J45.5, J46, J47, J60, J61, J62, J63, J64, J65, J66, J67, J68.4, J70.1, J70.3, J84.1, J84.9 |
| Renal Failure | I12.0, I13.1, N18, N19, N25.0, Z49.0, Z49.1, Z49.2, Z94.0, Z99.2 |
| Rheumatoid Arthritis, Collagen Vascular Diseases | L94.0, L94.1, L94.3, M05, M06, M08, M12.0, M12.3, M30, M31.0, M31.1, M31.2, M31.3, M32, M33, M34, M35, M45, M46.1, M46.8, M46.9 |
| Smoking | F17, I73.1, J41, J42, J43, J44, T65.2, Z71.6, Z72.0 |
| Solid Organ Transplantation | Z94.0, Z94.1, Z94.2, Z94.3, Z94.4 |

**eAppendix 1. Correlation Network of Comorbidities.**

Several underlying medical conditions (comorbidities) have been identified as increasing the risk for severe illness from the virus that causes COVID-19. Severe illness from COVID-19 is defined as hospitalization, admission to the ICU, intubation or mechanical ventilation, and death. The list of comorbidities associated with poor outcomes for patients infected with SARS-Cov-2 have evolved over time as we have learned much more about this disease throughout its course.

We quantified the strength of the comorbidities by calculating correlation coefficient associated with a pair of diseases using the following formula:

$$\Phi_{ij} = \frac{C_{ij}N - P_iP_j}{\sqrt{P_iP_j(N - P_i)(N - P_j)}}$$

where $C_{ij}$ corresponds to the number of patients affected by both comorbidities, $N$ is the total number of patients in the study population, and $P_i$ is the prevalence of the $i^{th}$ comorbidity. The distribution of $\Phi$ values represent all disease pairs where $C_{ij} > 0$. The $\Phi$ correlation is the Pearson's correlation for dichotomous variables (taking values of 0 or 1). We can determine the significant of $\Phi \neq 0$ by performing a $t$-test, according to the following formula:

$$t = \frac{\Phi\sqrt{n-2}}{\sqrt{1-\Phi^2}}$$

In the equation for $t$, $n$ corresponds to the number of observations used to calculate $\Phi$. We determined the level of significance of $t \geq 1.96$, significance at the 5% level. Correlation networks included in this study represent only those comorbidities with positive and significant correlation among each other. When two comorbidities co-occur more frequently than expected by chance, we have $\Phi > 0$. We quantified the comorbidity strength for the cohort over all, as well as, for subgroups according to whether or not a patient experienced a composite outcome (admitted to the ICU and/or died during hospitalization).

**eAppendix 2. Change-Point Detection Analysis of Hospitalizations, ICU Admissions, and In-Hospital Deaths.**

In this study we presented the characteristics of a large diverse cohort of patients with COVID-19 that were hospitalized in 5 UC Health hospitals in California. While the main aim of this study was to describe the COVID-19 experience and outcomes of these patients, we were able to also investigate the times series of hospitalizations, ICU admissions, and in-hospital death, mainly due to the fact that our case series expands from December 13, 2019, to January 6, 2021. Having access to an extensive times series, naturally allows for the study of change point analysis. In order to observe changes in trends in the number of hospitalizations, ICU admissions, and deaths, we smoothed the data with a moving average in a 7-day window (average time for a person to present symptoms and be captured by the health system). Data smoothing reveals points where case growth has accelerated and slowed. Further identification of time points during the study period of multiple change points within the time series can very insightful to improve our understanding of the impact of this pandemic in UC Health hospitals.

In this study we performed a multiple change point detection using the binary segmentation algorithm (ref), which aims to identify multiple change points by minimizing the following cost function $\mathrm{C}$:

$$\sum_{i=1}^{m+1} [\mathrm{C}(y_{\tau_i - 1+1):\tau_i}))] + \beta f(m)$$

Where $\mathrm{C}$ is the cost function for segment (e.g., negative log-likelihood) and $\beta f(m)$ is a penalty guard against over fitting. The *changepoint* package that we implemented in *R* software implements used binary segmentation as the algorithm (Edwards and Cavalli-Sforza 1965).

Binary segmentation applies first a single changepoint test statistic to the entire time series data, if a changepoint is identified, then the data is split into two at the changepoint location determined in the first phase. This process is repeated on two new data states, before and after the change was identified. If changepoints are identified in either of the two new data sets, they are split further. This process is repeated until no changepoints are found sin any parts of the times series data. This procedure is an approximate minimization of the cost function C and it is computationally fast compared to other algorithms aimed to minimize the cost function.

**eFigure 1. Change Points (means) in the number of patients admitted to the intensive care unit, and in-hospital deaths. Data smoothing reveals and change point analysis revealed that hospital admissions and patients treated in the ICU changed on the same dates, while changepoints for deaths occurred approximately a week after hospitalizations and ICU admissions.**

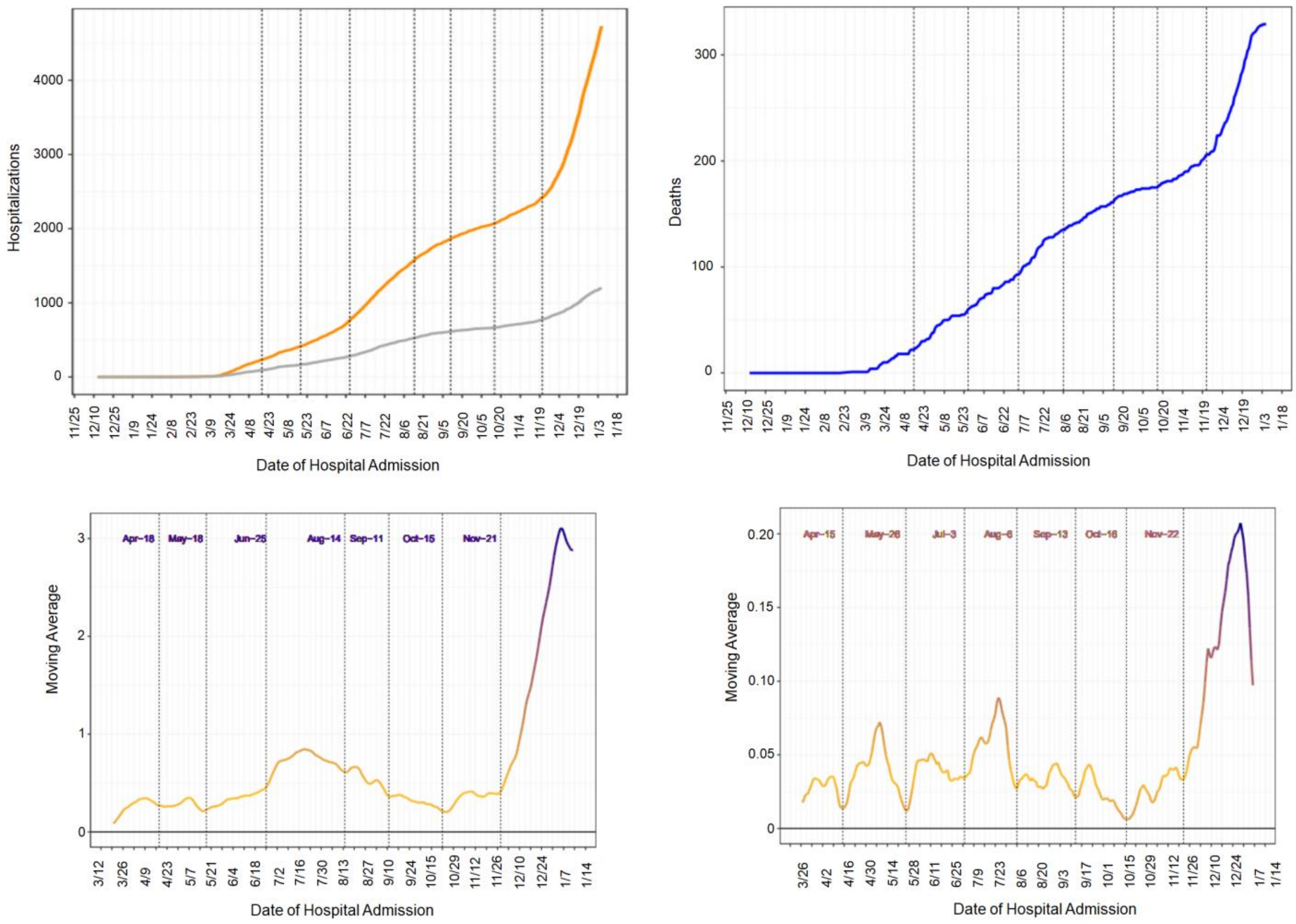
